# Leveraging Serosurveillance and Postmortem Surveillance to Quantify the Impact of COVID-19 in Africa

**DOI:** 10.1101/2022.07.03.22277196

**Authors:** Nicole E. Kogan, Shae Gantt, David Swerdlow, Cécile Viboud, Muhammed Semakula, Marc Lipsitch, Mauricio Santillana

## Abstract

**Background:** The COVID-19 pandemic has had a devastating impact on global health, the magnitude of which appears to differ intercontinentally: for example, reports suggest 271,900 per million people have been infected in Europe versus 8,800 per million people in Africa. While Africa is the second largest continent by population, its reported COVID-19 cases comprise <3% of global cases. Although social, environmental, and environmental explanations have been proposed to clarify this discrepancy, systematic infection underascertainment may be equally responsible.

**Methods:** We seek to quantify magnitude of underascertainment in COVID-19’s cumulative incidence in Africa. Using serosurveillance and postmortem surveillance, we constructed multiplicative factors estimating ratios of true infections to reported cases in African nations since March 2020.

**Results:** Multiplicative factors derived from serology data – in a subset of 12 nations – suggested a range of COVID-19 reporting rates, from 1 in 630 infections reported in Kenya (May 2020) to 1 in 15 infections reported in South Africa (November 2021). The largest multiplicative factor, 3,795, corresponded to Malawi (June 2020), suggesting <0.05% of infections captured. A similar set of multiplicative factors for all nations derived from postmortem data points toward the same conclusion: reported COVID-19 cases are unrepresentative of true infections, suggesting a key reason for low case burden in many African nations is significant underdetection and underreporting.

**Conclusions:** While estimating COVID-19’s exact burden is challenging, the multiplicative factors we present provide incidence curves reflecting likely-to-worst-case ranges of infection. Our results stress the need for expansive surveillance to allocate resources in areas experiencing severe discrepancies between reported cases, projected infections, and deaths.

**Summary:** Here we present a range of estimates quantifying the extent of underascertainment of COVID-19 cumulative incidence in Africa. These estimates, constructed from serology and mortality data, suggest that systematic underdetection and underreporting may be contributing to the seemingly low burden of COVID-19 reported in Africa.

## Introduction

In just over 2 years, coronavirus disease 19 (COVID-19) has led to a reported 545 million cases and 6.3 million deaths globally [1]. Asia, North America, South America, and Europe have respectively contributed 156 million, 103 million, 59 million, and 204 million cases to this toll (roughly 33,400, 172,100, 136,400, and 271,900 cases per million population), as well as over 1.3 million deaths each, whereas reported COVID-19 morbidity and mortality have appeared considerably lower in Africa: 12 million cases (8,800 cases per million population) and 254,900 deaths [1, 2].

This observation has led some to conclude that the pandemic has “spared” Africa and parts of Asia [3]. Although several explanations have been proposed – including strong government response, young age structure, sparse population density, competing comorbidities, and climate effects – the World Health Organization (WHO) has instead posited that systematic underascertainment explains the continental difference in reporting [3-6]. Leveraging the COVID-19 prevalence calculator developed by the global health organization Resolve to Save Lives, the WHO has further estimated that only 1 in 7 COVID-19 cases in Africa have been detected, compared to 1 in 4 COVID-19 cases in the United States [4, 7, 8]. The WHO’s hypothesis has been bolstered by several recent national and sub-national studies [3, 9]. However, the variability in underascertainment reported by these studies motivates the need for practicable methods that can triangulate between disparate data sources to provide credible estimates of African nation-specific COVID-19 infections. While the exact number of infections can never be known, producing a range of estimates reflective of likely- and worst-case scenarios is still valuable in understanding the burden of disease. Here we propose 2 such methods that approximate the magnitude of COVID-19 cumulative incidence in Africa, the first relying on serosurveillance data and the second relying on postmortem surveillance data.

## Methods

### Motivation for Serosurveillance

National population-based serosurveys furnish cross-sectional estimates of cumulative incidence of infection by SARS-CoV-2 in quantifying the proportion of a population with SARS-CoV-2 antibodies (“seroprevalence”). Most relevant for infection are the antibody isotypes IgA, IgG, and IgM, which can be further classified based on their specificity for SARS-CoV-2 proteins spike (S), receptor binding domain (RBD), or nucleocapsid (N) [10, 11]. Antibody specificity can also help differentiate between antibody production due to natural infection versus vaccination (coverage for which is still low in Africa), where the latter is observed to induce production of anti-S and anti-RBD – but not anti-N – antibodies [11, 12, 13].

### Inferring Infection Underascertainment: Analysis of Serosurveillance Data

Using SeroTracker, a global SARS-CoV-2 seroprevalence database, we identified 29 national-level serosurveys collected over the course of the pandemic across 12 African nations: Cape Verde, Côte d’Ivoire, Egypt, Ethiopia, Gabon, Ghana, Kenya, Malawi, Senegal, Sierra Leone, South Africa, and Zambia [14].

Table 1 shows the sample size and seroprevalence for each such serosurvey. SeroTracker not only furnishes a point estimate and a 95% confidence interval for seroprevalence at a given time point during the pandemic but also provides metadata for each serosurvey. This metadata includes sampling characteristics – method, location, timeframe, corresponding publication, Joanna Briggs Institute-based bias risk, population group, and population age – and serosurvey characteristics – manufacturer, test type, detected isotype, sensitivity, and specificity [15].

**Table 1:** Summary of Serosurveillance Study Metadata from SeroTracker^*^.

| **Nation** | **# Tested** | **Seroprevalence**<br>**(95% CI)** | **Sampling Period**<br>**(MM.DD.YY)** | **Population** | **Ages** |
| --- | --- | --- | --- | --- | --- |
| Cape Verde | 5,348 | 0.4% | 06.26.20-07.04.20 | Household and community | Multiple groups |
| Côte d'Ivoire | 1,687 | 25.1% | 07.15.20-10.13.20 | Essential non-HCW | 18-64 years |
| Egypt | 1,260<br>82 | 34.8%<br>46.3% | 10.01.20-10.31.20<br>10.15.20-12.15.20 | Household and community<br>HCW and caregivers | Multiple groups<br>18-64 years |
| Ethiopia | 16,932<br>1,997 | 3.5% (3.2%-3.8%)<br>39.8% (37.4%-41.7%) | 06.24.20-07.08.20<br>12.15.20-02.15.21 | Household and community<br>HCW and caregivers | Multiple groups<br>18-64 years |
| Gabon | 319 | 55.0% | 03.15.20-08.15.20 | COVID-19 patient contacts | Multiple groups |
| Ghana | 1,305<br>395<br>393<br>636 | 19.0%<br>15.2%<br>9.7%<br>22.8% | 07.27.20-09.14.20<br>10.01.20-10.31.20<br>12.01.20-12.31.20<br>02.01.21-02.28.21 | Multiple general populations | Multiple groups |
| Kenya | 830<br>684<br>9,922<br>3,018<br>176<br>176 | 42.3% (38.4%-46.3%)<br>20.8% (17.5%-24.4%)<br>7.9% (6.7%-9.0%)<br>48.5% (45.2%-52.1%)<br>83.0%<br>2.3% | 09.30.20-10.23.20<br>07.30.20-12.04.20<br>04.30.20-09.30.20<br>01.03.20-03.15.21<br>08.01.21-08.31.21<br>05.01.20-05.31.20 | Essential non-HCW<br>HCW and caregivers<br>Blood donors<br>Blood donors<br>Blood donors<br>Blood donors | 18-64 years<br>18-64 years<br>18-64 years<br>Multiple groups<br>18-64 years<br>18-64 years |
| Malawi | 500<br>5,085 | 12.3% (8.2%-16.5%)<br>27.6% | 05.22.20-06.19.20<br>01.15.20-07.15.21 | HCW and caregivers<br>Blood donors | 18-64 years<br>Multiple groups |
| Senegal | 303<br>1,422 | 21.2% (16.7%-26.1%)<br>28.4% (26.1%-30.8%) | 06.15.20-09.15.20<br>10.25.20-11.26.20 | Non-COVID-19 patients<br>Household and community | Multiple groups |
| Sierra Leone | 1,893 | 2.6% (1.9%-3.4%) | 03.01.21-03.31.21 | Household and community | Multiple groups |
| South Africa | 927<br>16,762<br>7,577<br>3,395 | 35.8%<br>47.4% (46.2%-48.6%)<br>45.2% (43.7%-46.7%)<br>71.1% (68.8%-73.5%) | 11.23.20-01.31.21<br>01.15.21-05.15.21<br>11.15.20-04.15.21<br>11.08.21-11.12.21 | Household and community<br>Blood donors<br>Multiple general populations<br>Blood donors | Multiple groups |
| Zambia | 2,704<br>575<br>1,657 | 2.1% (1.1% - 3.1%)<br>2.2% (0.5% - 3.9%)<br>8.2% (5.1% - 11.4%) | 07.04.20-07.27.20<br>07.02.20-07.31.20<br>07.02.20-07.31.20 | Household and community<br>HCW and caregivers<br>Residual sera | Multiple groups |

\* *Abbreviations: HCW, healthcare workers; “multiple groups”, at least a 30% overlap within* *another age group*

For each of the 12 African nations, seroprevalences were compared to the proportion of that nation’s population reported to have COVID-19. The latter proportion was obtained by dividing reported cumulative cases (Our World in Data) at the end of the serosurvey sampling period by the corresponding nation’s population in 2020 (World Bank) or 2021 (United Nations) [16, 17]. Hereafter, we refer to the ratio of seroprevalence to this proportion as a “multiplicative factor.” This multiplicative factor-based framework extends the work of Angulo et al. (2021), who previously formulated a procedure to estimate COVID-19 infections (both asymptomatic and symptomatic), hospitalizations, and deaths in the United States using serosurveys [9].

A multiplicative factor of 1 would suggest complete concordance in population positivity for COVID-19 between serology reports and case reports – equivalently, no apparent underascertainment. In contrast, a multiplicative factor greater than 1 would suggest that serology-derived estimates of infection incidence exceed reported cases, reflective of a “hidden morbidity” – of apparent underascertainment. Multiplicative factors are generally expected to be greater than 1 due to the large number of asymptomatic COVID-19 infections that go undetected in nearly all settings, the insensitivity of tests, the failure to report positive tests, and/or the absence of testing among certain populations. Applying multiplicative factors to nation-specific epidemic curves produces scaled curves that estimate infection incidence while maintaining local-in-time trends (Figure 1). To limit overextrapolation, these scaled curves were only projected to the nearest half-year. Although there is a 1-to 2-week delay between infection and antibody positivity, lagged reporting of infections counteracts the effect of this delay [18,19].

**Figure 1:**
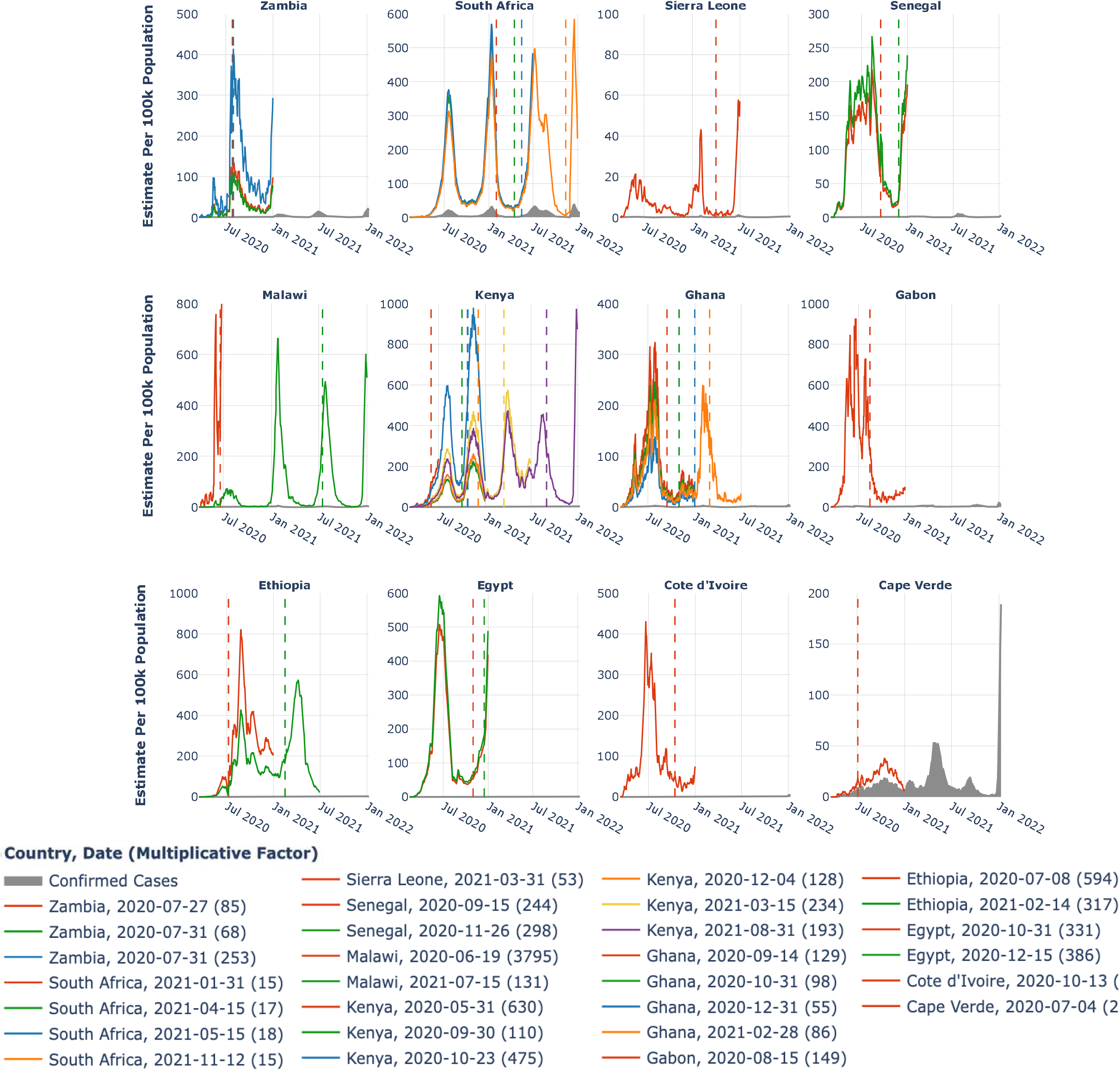
Confirmed COVID-19 Cases and Serology-Based Infection Estimates for 12 African nations. Reported COVID-19 cases (gray) versus seroprevalence-derived COVID-19 infections (red, green, blue, orange, yellow, purple) per 100,000 population for 12 African nations. Each non-gray curve is created through scaling the corresponding gray curve by a multiplicative factor, parenthetically indicated in the legend, with scaling performed to the nearest half-year to reduce overextrapolation of results. Color order corresponds to chronological temporal order (red: earliest, purple: latest) of serosurvey. Vertical dotted lines represent the last date of each serosurvey sampling period.

### Inferring Infection Underascertainment: Analysis of Postmortem Surveillance Data

To create another set of multiplicative factors, we made use of 2 Zambian postmortem surveillance studies that employed quantitative reverse transcription PCR to detect COVID-19 in decedents: Mwananyanda et al.’s (2021) postmortem surveillance study, as well as its preprinted (not yet peer-reviewed) follow-up from Gill et al. (2022) [20, 21]. Both studies were performed in a sample from Zambia’s capital, Lusaka, with the first conducted between June and October 2020 following the first wave of infections and the second conducted between January and June 2021 following the second wave of infections [22].

Prior to these studies, fewer than 10% of deaths attributed to or related to COVID-19 in Lusaka were identified in life with antemortem testing [19]. In contrast, the 2 postmortem studies found that 58 of 364 (15.9%) decedents in the first and 358 of 1,116 (32%) decedents in the second showed PCR positivity, suggesting marked underascertainment of COVID-19 cases and deaths. With much of Africa, including Zambia, identified as “high risk” on the Infectious Disease Vulnerability Index, it is not improbable that these percentages reflect reality in other African nations [23]. Under this assumption, we multiplied 15.9% and 32% by each African nation’s estimated all-cause mortality in 2020 and in 2021, respectively, to obtain the expected number of year-end deaths with COVID-19 (Supplementary Figure 1) [16, 17]. Each of these expectations was then divided by infection fatality ratios (IFRs) – defined as a comparison of infections to deaths due to infection – obtained using different parametric methods by Sorensen et al. (2022) and Onovo et al. at multiple time points during the pandemic [24, 25]. The result is the expected number of year-end COVID-19 infections (Figure 2, Supplementary Figure 2), which can be compared to reported cumulative COVID-19 cases in 2020 and in 2021 to produce another set of multiplicative factors that expand the range of credible infection estimates. These multiplicative factors are visualized in Figure 3.

**Figure 2:**
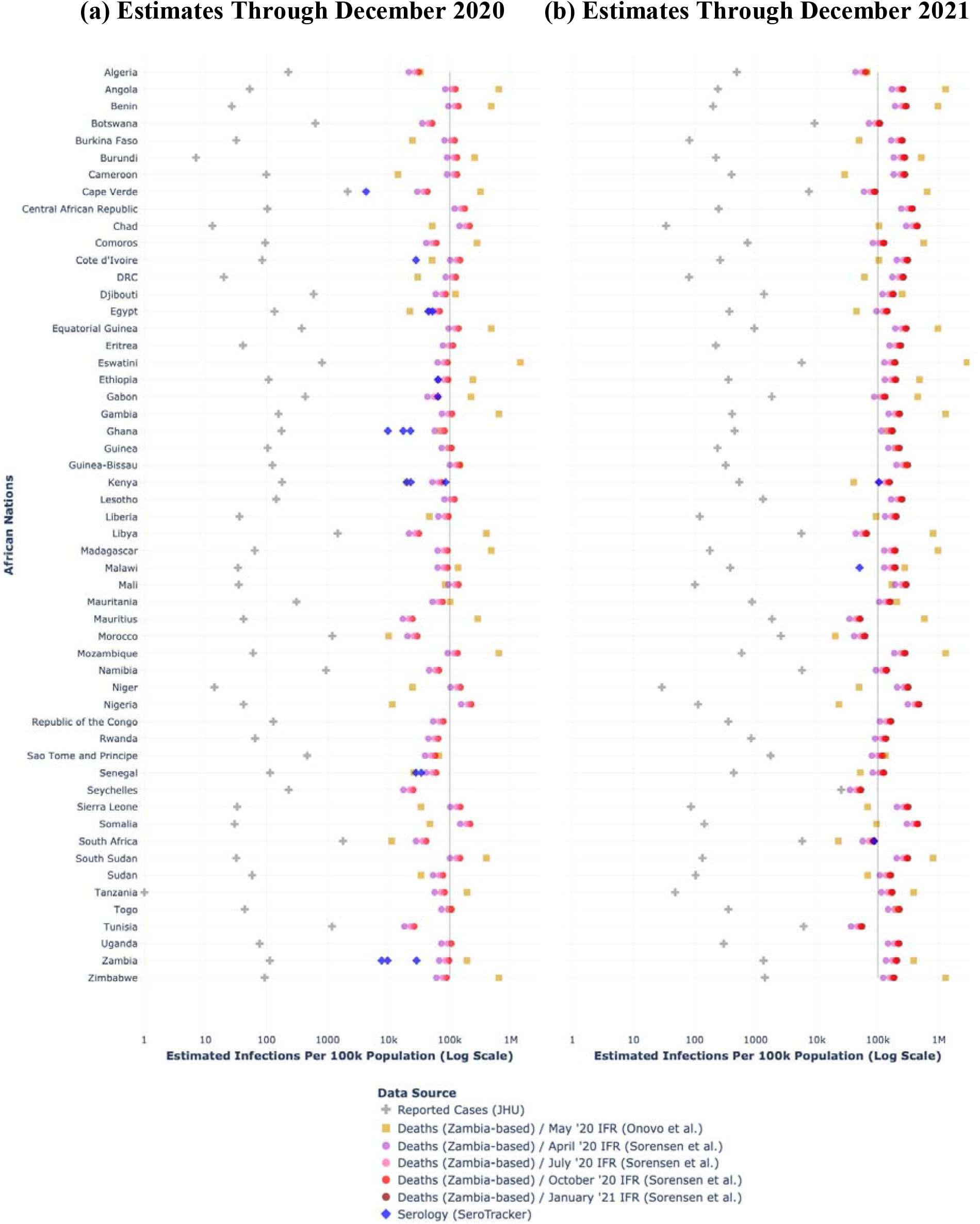
Estimated COVID-19 Infections Using Serosurveillance and Postmortem Surveillance in Africa. Comparison of reported COVID-19 cases, cumulative seroprevalence-derived COVID-19 infections (available for 12 nations), and cumulative postmortem-derived COVID-19 infections (Onovo et al. available for 44 nations, Sorensen et al. available for 54 nations) per 100,000 population in Africa. The solid black line in each subplot represents 100,000 infections per 100,000 population, implying that an entire population has been infected. **(a)** represents infection estimates through December 31, 2020, and it draws from serosurveys taken between July 1, 2020 and December 31, 2020 as well as from postmortem surveys detailed in Mwananyanda et al. (2021) to derive Zambia-based COVID-19 deaths. **(b)** represents infection estimates through December 31, 2021, and it draws from serosurveys taken between July 1, 2021 and December 31, 2021 as well as from postmortem surveys detailed in Gill et al. (2022) to derive Zambia-based COVID-19 deaths.

**Figure 3:**
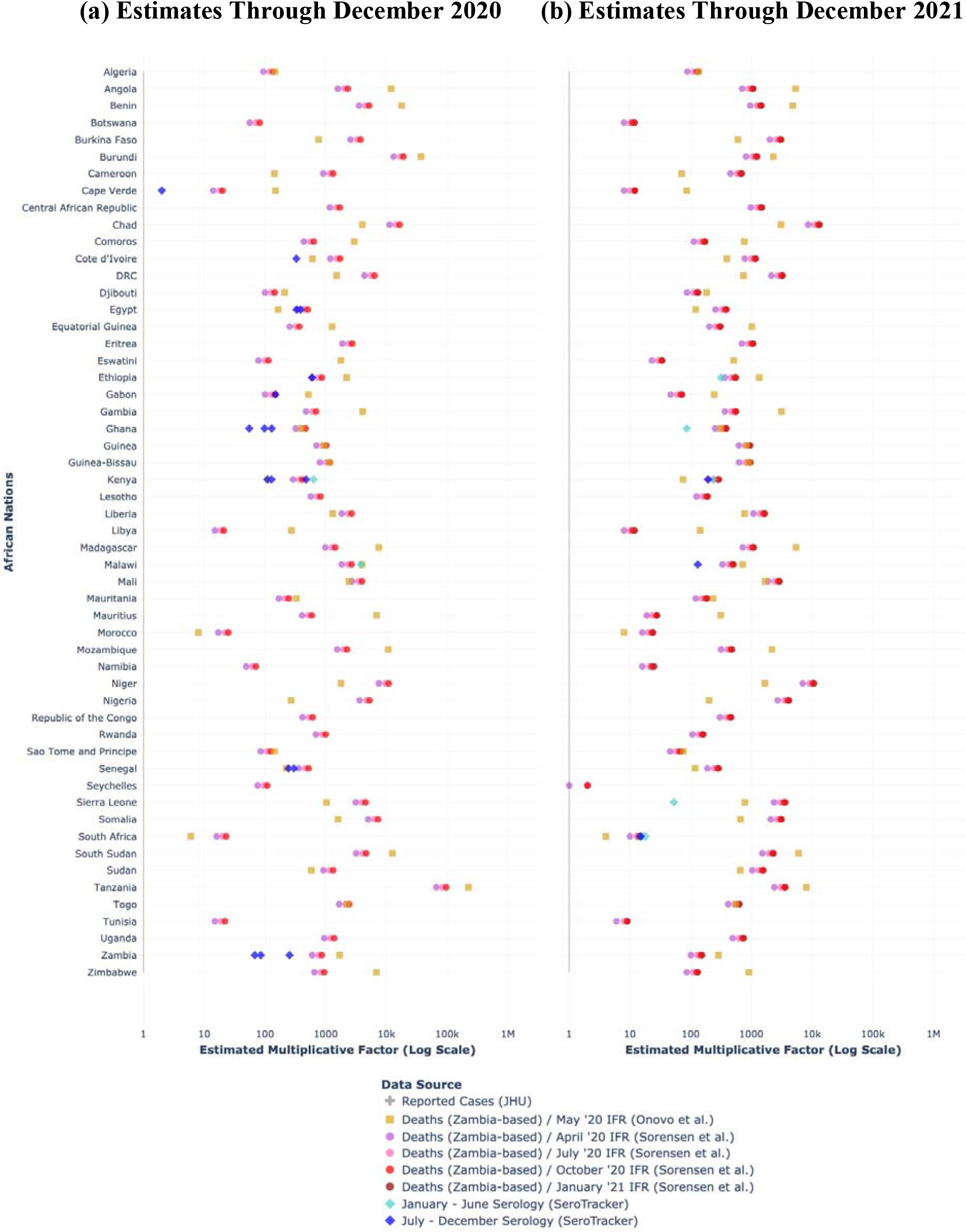
Multiplicative Factors Needed to Estimate True Infections from Confirmed Cases of COVID-19 in Africa. Comparison of multiplicative factors summarizing ratio of seroprevalence-derived COVID-19 infections (available for 12 nations) and postmortem-derived COVID-19 infections (Onovo et al. available for 44 nations, Sorensen et al. available for 54 nations) to reported cases in Africa. **(a)** represents ratio estimates through December 31, 2020, and it leverages postmortem surveys detailed in Mwananyanda et al. (2021) to derive Zambia-based COVID-19 deaths. **(b)** represents ratio estimates through December 31, 2021, and it leverages postmortem surveys detailed in Gill et al. (2022) to derive Zambia-based COVID-19 deaths.

Note that when discussing these data, we intentionally use the phrase “deaths with COVID-19” instead of “deaths from COVID-19” because inferring causality is a challenge in postmortem surveillance. Even so, the United States Centers for Disease Control and Prevention have issued guidance that COVID-19 should be assumed the underlying cause of death if SARS-CoV-2 is detected [26].

## Results

### Serology Multiplicative Factors

The available serosurveys result in multiplicative factors that are illustrative of systematic underdetection and underreporting (Figure 1). Multiplicative factors ranged in magnitude from 2 (Cape Verde, based on data from July 2020) to 3,795 (Malawi, based on June 2020 data), implying that 1 in 2 and 1 in 3,795 infections were respectively detected in these nations early in the pandemic progression. More than half of these multiplicative factors were at the “hundreds” order of magnitude, which facilitates discernment of 4 distinct COVID-19 “waves” previously obscured by the low amplitude peaks of confirmed cases (Figure 1) [22].

Additionally, Kenya, for which the largest number of national-level serosurveys were available (6, spanning April 2020 to September 2021), produced multiplicative factors ranging from 110 in December 2020 to 630 in May 2020. This observation of declining multiplicative factors as the pandemic progressed is echoed in data for Malawi, Ghana, Ethiopia, and South Africa, which may reflect incremental improvements in testing capacity.

### Inter-Method Comparison: Estimates of Infection

Figure 2 provides an inter-method comparison of estimated infections (per 100,000 population). For many nations, the upper bound estimate of COVID-19 infection burden was produced by the estimator shown in gold, which couples postmortem surveillance data from either Mwananyanda et al. (2021) or Gill et al. (2022) with IFR data from Onovo et al. (2021). The large magnitude of the gold estimates is less likely a byproduct of the timing of the Onovo et al. (2021) IFR estimates – May 2020 – and more likely a byproduct of differences in IFR estimation methodologies, as the [even earlier] April 2020 IFR estimates from Sorensen et al. (2022) generated infection estimates that are generally lower by several orders of magnitude.

The lower bound estimate of COVID-19 infection burden was produced by the estimator shown in blue, which leverages serosurveillance data. Additionally, infection estimates for most nations cluster around 100,000 infections per 100,000 population. This signals that the majority of individuals in most populations may have already experienced infection at least once.

### Inter-Method Comparison: Multiplicative Factors

All African nations yielded multiplicative factors exceeding 1 for the serosurveillance method and the postmortem surveillance method (Figure 3), which is indicative of divergence between “ground truth” infections and reported cases.

Also notable in Figure 3 is concordance among the multiplicative factors calculated using IFRs from Sorensen et al. (2022), as observed in the clustering of red-shaded markers. Of these, the purple marker, which reflects IFRs from April 2020, corresponded to the smallest magnitude multiplicative factor, whereas the maroon marker, which reflects IFRs from January 2021, corresponded to the largest magnitude multiplicative factor.

Multiple North African nations (Tunisia, Morocco, Libya), island nations (Cape Verde, Seychelles), Botswana, and South Africa yielded the smallest magnitude multiplicative factors (<20). The similarly small magnitude across these nations is notable in that South Africa’s established indicator- and event-based disease surveillance system and Cape Verde’s and Seychelles’ geographic isolation contrasts with weakened public health infrastructure in the Middle East and North Africa (MENA) region, as many MENA nations have grappled with a paucity of resources to adequately chronicle COVID-19 [27].

Finally, the change in x-axis scale from subplot (a) to subplot (b) also reveals an order-of-magnitude decrease in multiplicative factors from 2020 to 2021, perhaps coincident with improved COVID-19 testing infrastructure and increased implementation of pharmaceutical and non-pharmaceutical interventions over the pandemic’s progression.

### Assessment of Vulnerabilities

One explanation for underreporting and underdetection is tied to health, economic, and social vulnerabilities. Lewis et al. (2022) describe analysis of these vulnerabilities as an unmet need [6]. These vulnerabilities include food insecurity, poverty, and noncommunicable disease comorbidity, among others [28].

One tool to assess these vulnerabilities is Surgo Ventures’ Africa COVID-19 Community Vulnerability Index (CCVI) [29]. Across and within 756 regions in 48 African nations, CCVI specifically encodes 7 dimensions of vulnerability, which can together or individually contribute to heterogeneous attenuation of disease signal: age, epidemiology, fragility, health system, population density, socioeconomics, and transport and housing availability. These dimensions are quantified on a Likert-like scale as “very low”, “low”, “moderate”, “high”, or “very high”. We limit our focus to epidemiological factors (non-communicable diseases, non-COVID-19 infectious diseases) and health system factors (strength, capacity, and access).

In Supplementary Figure 3, we ranked African nations by variance in their respective multiplicative factors and compared this to nation-specific health system, epidemiological, and overall vulnerabilities (as quantified by CCVI). By way of example, South Africa exhibited not only low variance in its multiplicative factor estimates but also low “health system” vulnerability, the combination of which seems to reflect more robust surveillance infrastructure.

Calculating the Spearman Rank correlation between the ascending variances for 2020 and the correspondingly ordered “health system” vulnerability index values yields a coefficient of 0.44 and a corresponding p-value of 0.001. At a prespecified a-level of 0.05, we reject the null hypothesis that health system vulnerability and variability in infection estimates are uncorrelated and conclude that there exists some evidence to suggest a correlation. While this comparison does not establish a definitive relationship, on average, nations with lower inter-method variances have a lower degree of vulnerability compared to nations with higher inter-method variances.

## Discussion

Although underascertainment of COVID-19 remains a global challenge, the degree to which COVID-19 cumulative incidence is underdetected and underreported in Africa appears higher than in other continents. By comparison, for example, multiplicative factors were previously estimated to be between roughly 5 and 50 in North America [30, 31]. Both the serosurveillance-and postmortem surveillance-based estimates we present here suggest that COVID-19 infections are tens-to hundreds-fold greater than their reported counterparts.

Our process is complementary to a preprinted (not yet peer-reviewed) meta-analysis from Lewis et al. (2022), which also seeks to quantify the divergence between seroprevalence-derived infections and reported cases in Africa [6]. However, the meta-analysis relied predominantly on local-level serosurveys for inference, where local-level serosurveys are primarily obtained in urban settings. The national-level serosurveys we leverage are intended to increase generalizability of results (i.e., without limiting inference to cities). Coarser resolution – as well as not restricting serosurveys under study to those that strictly align with SEROPREV protocol criteria – also means that our analysis includes areas un- or underrepresented in Lewis et al.’s work, such as Egypt and Cape Verde [32].

We additionally do not limit the scope of our analysis to serosurveillance data, validating our results with postmortem surveillance data. Postmortem surveillance data has the added advantage of facilitating direct estimation of COVID-19-attributable deaths, where deaths have also exhibited underascertainment globally [33]. As Mwananyanda et al. (2021) and Gill et al. (2022) show, in Lusaka, Zambia, fewer than 10% of those who died with or from COVID-19 were tested for COVID-19 prior to death [19]. Supplementary Figure 1 in part captures this underreporting and underdetection of deaths, comparing reported COVID-19-attributable deaths in 2020 and 2021 to Zambia- and excess mortality-based deaths estimates [34]. Although several nations exhibit negative excess mortalities (e.g., Seychelles and Mauritius), which would seem to suggest that the COVID-19 pandemic led to a reduction in deaths, inflection points in the curves at the time of first recorded COVID-19 deaths illustrates that there was instead an existing negative trend in all-cause deaths that COVID-19 reversed [35].

The results from our multiplicative framework are concordant with other methods. For example, a stochastic [multi-strain] compartmental national epidemic model for South Africa in a preprinted (not yet peer-reviewed) analysis from Gozzi et al. (2022) suggests that South Africa’s surveillance system could detect 1 in 16 infections between May 2021 and November 2021, compared to our estimates of multiplicative factors between 10 and 20 for the same timeframe [36]. Additionally, using a global metapopulation epidemic model, Davis et al. (2021) recovered infection attack rates around 1% for several African nations early in the pandemic’s progression, further underscoring the discrepancy between recorded, reported cases and true infections [31].

A limitation of our approach is that the magnitude of multiplicative factors derived from the serosurveillance method may be associated with serosurvey-specific risk of bias. Supplementary Figure 4 explores this association by stratifying the roughly 30 serosurveys by their bias risk, as measured by the Joanna Briggs Institute Checklist for Prevalence Studies (JBI). JBI is a validated critical appraisal tool that encompasses 9 dimensions of assessment, which SeroTracker uses to classify each serosurvey [14, 15]. Ultimately, we found that although the “high bias” stratum corresponded to the highest median multiplicative factor (161) and smallest dispersion compared to the “low bias” stratum and “medium bias” stratum (128 and 110), the overlapping nature of the box plots suggests that this difference is qualitative at best and, therefore, that multiplicative factors of larger magnitude may not be any more biased than multiplicative factors of smaller magnitudes. Furthermore, the propensity for bias does not translate to the direction of bias: simply because a serosurvey is classified as biased does not indicate that resulting seroprevalence is lower or higher than true prevalence. Therefore, we cannot establish that higher multiplicative factors are indeed a result of higher bias.

A different source of bias in our serosurveillance method relates to origins of seropositivity, specifically differentiating between seropositivity from vaccination or from infection. However, this is less likely to be of concern given the low vaccination rate of 10% in Africa for the 2020 – 2021 period of analysis [13]. Even so, about one third of tests chronicled by SeroTracker measure IgG (about half are anti-S), one third measure both IgM and IgG, and one third measure total antibody [14]. Additionally, we do not explicitly account for waning immunity and resulting reinfection, which may have contributed to certain nations exhibiting estimates of infection that exceed the total population (i.e., any points in Figure 2 and Supplementary Figure 2 exceeding 100,000 cases per 100,000 population). Relatedly, we cannot rule out underestimation of “ground truth” disease burden due to seroreversion, the phenomenon in which antibody levels decline over time to a level below the cutoff for seropositivity, resulting in an underestimation of cumulative incidence [37]. Seroreversion and reinfection would each lead to underestimates of cumulative incidence, and thus underestimates of the multiplicative factor between reported cases and true infections. This further underscores the narrative that reported cases do not adequately capture the burden of infection.

Another important feature of serosurveys is the populations from which they sample. These populations often include healthcare workers, blood donors, and the elderly, who may be differentially exposed to SARS-CoV-2. Our analysis does not consider these characteristics of the sample frame and, therefore, may overestimate cumulative incidence of COVID-19 if the sampled individuals indeed have higher cumulative incidence than the underlying population. In Malawi, for example, the thousands order of magnitude for the June 2020 multiplicative factor could be in part attributable to an especially unrepresentative sample of healthcare workers and caregivers. Given the scarcity of reported infections stratified on serum donor profiles, one possibility for mitigating this bias would entail the extrapolation of seroprevalence to the general population using weights that account for demographic differences. The United States Centers for Disease Control and Prevention recently published such a reweighting protocol specific to blood donors [38]. Another possibility would be to assess intra-nation variability of serology estimates, as nations like Cameroon and South Africa have started releasing local-level serosurveys to the public, and to employ these estimates in a sensitivity analysis for multiplicative factor heterogeneity [39].

An additional consideration for contextualizing our results is that our methods combine multiple timeframes to create time-invariant multiplicative factors. For example, the serosurveillance studies from SeroTracker cover multiple dates between June 2020 and November 2021, whereas postmortem surveillance studies reflect either the time period of June 2020 to October 2020 or January 2021 to July 2021. It follows that multiplicative factors estimated from early pandemic stages may not be as representative of disease dynamics seen in later pandemic stages from SARS-CoV-2 strain-specific effects (e.g., Delta and Omicron). However, the cross-sectional nature of serosurveys suggests that cumulative estimates of infection, such as those presented in our analysis, may be more reliable than conversions to weekly estimates of infection.

Finally, in calculating the estimated number of infections using serosurveillance and postmortem surveillance data, we employed 2020 and 2021 national population size estimates from the World Bank and the United Nations [16, 17]. However, these estimates were the result of projections prior to the COVID-19 pandemic, which would have the effect of overestimating population size due to unaccounted for deaths. To quantify the impact of this compounding uncertainty, we performed a sensitivity analysis for 2021 infections and deaths by subtracting the number of COVID-19-attributable deaths in 2020 from the projected population size in 2021. The percent difference between unadjusted infections and adjusted infections was less than 0.5% for all African nations, a result summarized in Supplementary Table 1, which suggests that the projected population sizes without adjustment are sufficient for analysis.

Asymptomatic infections and variable rates of testing for COVID-19 in Africa, as well as globally, have made it particularly challenging to ascertain the true burden of the pandemic from data, as reflected in the range of multiplicative factors from our analysis (1 in 2 cases reported for Cape Verde, 1 in 3,795 cases reported in Malawi) [40, 41]. While previous studies have sought to estimate this systematic underascertainment in resource-limited settings, they have relied on a single data stream to make inferences. The innovation of our approach lies in leveraging not only serosurveillance data (antibody testing) but also postmortem surveillance data (PCR testing) to characterize COVID-19 in Africa. Concordance between these methods, when taken with other sources of epidemiological data, such as syndromic surveillance and digital surveillance [19, 42] can be leveraged in 2 ways: (1) to help inform allocation of resources to areas exhibiting noticeable divergence in reported infections and recalibrated infections; and (2) to help inform initiatives that can bolster surveillance infrastructure.

## Data Availability

All data produced in the present study are available upon reasonable request to the authors.

https://serotracker.com/en/Explore

## Acknowledgements

We thank Bethany Hedt-Gauthier (Harvard T.H. Chan School of Public Health) for her thorough review of the manuscript and for her facilitation of international collaboration in this work. We also thank Amit Srivastava (Pfizer Inc.) for his invaluable industry perspective.

