## Supplement for "Leveraging Serosurveillance and Postmortem Surveillance to Quantify the Impact of COVID-19 in Africa"

**Supplementary Figure 1**

**(a) Estimates Through December 2020	(b) Estimates Through December 2021**

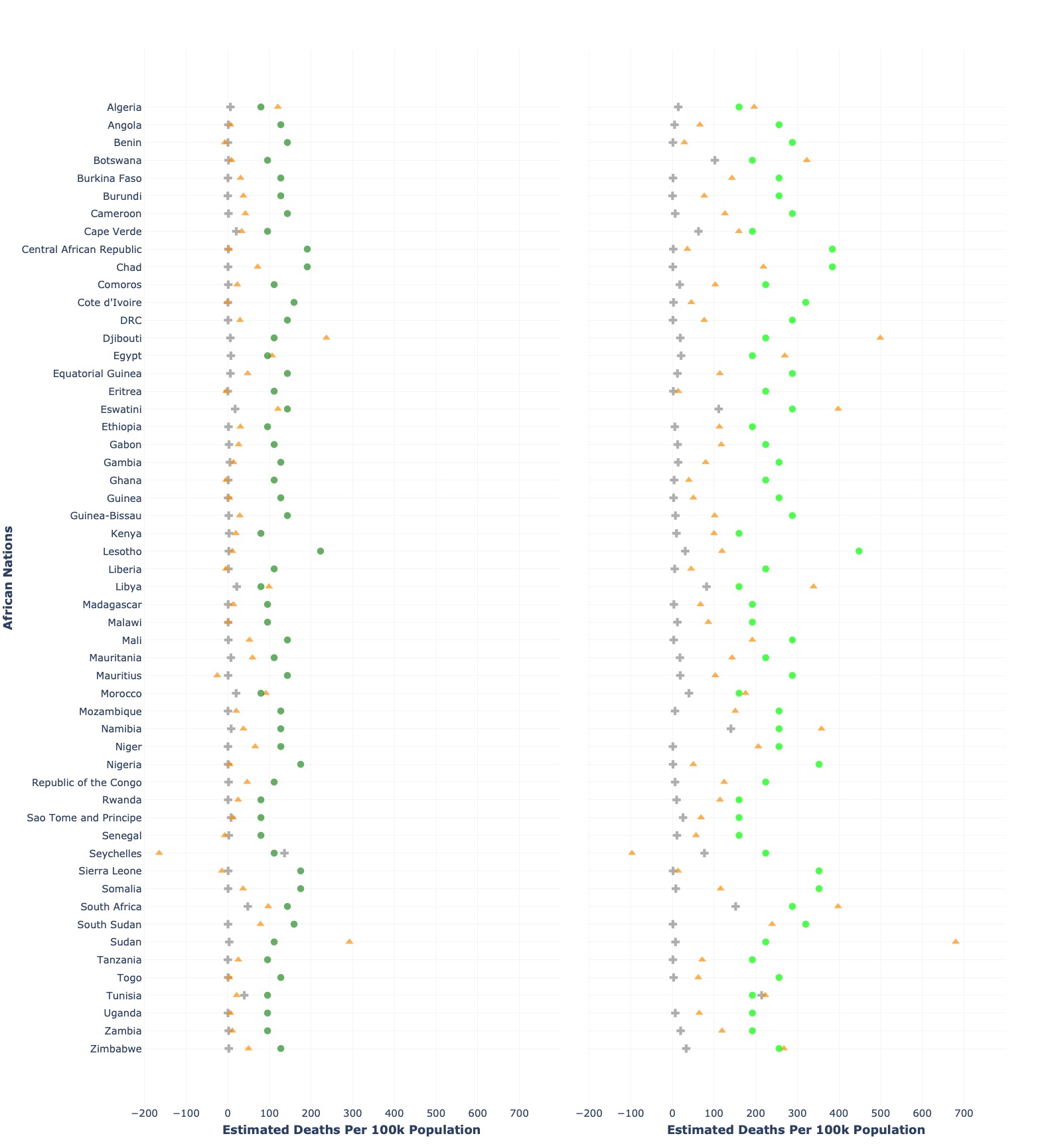

*Estimated number of deaths with COVID-19 in Africa (n = 54 countries) per 100,000 population. Gold-colored markers represent estimated deaths derived from excess mortality (The Economist), dark green-colored markers represent estimated deaths derived from Mwananyanda et al. (2021), and light green-colored markers represent estimated deaths derived from Gill et al. (2022).*

**Supplementary Figure 2**

**(a) Estimates Through June 2020	(b) Estimates Through June 2021**

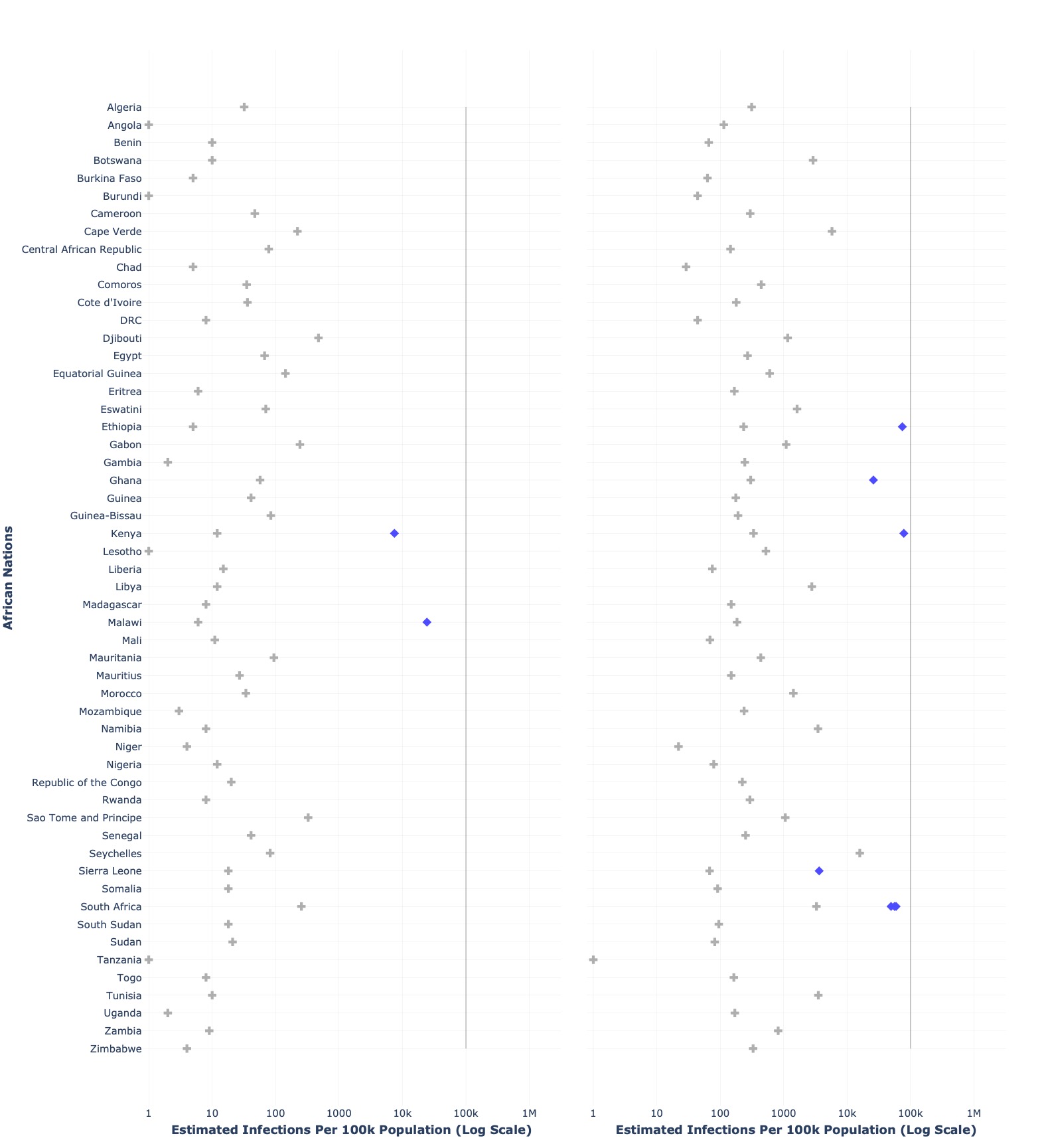

*Comparison of reported COVID-19 cases, cumulative seroprevalence-derived COVID-19 infections (available for 12 nations), and cumulative postmortem-derived COVID-19 infections (Onovo et al. available for 44 nations, Sorensen et al. available for 54 nations) per 100,000 population in Africa. The solid black line in each subplot represents 100,000 infections per 100,000 population, implying that an entire population has been infected. Since mortality is reported annually and not semi-annually, only serology-derived estimates are shown.* ***(a)*** *represents infection estimates through June 30, 2020, and it draws from serosurveys taken before this date* ***(b)*** *represents infection estimates through June 30, 2021, and it draws from serosurveys taken between January 1, 2021 and June 30, 2021.*

**Supplementary Figure 3**

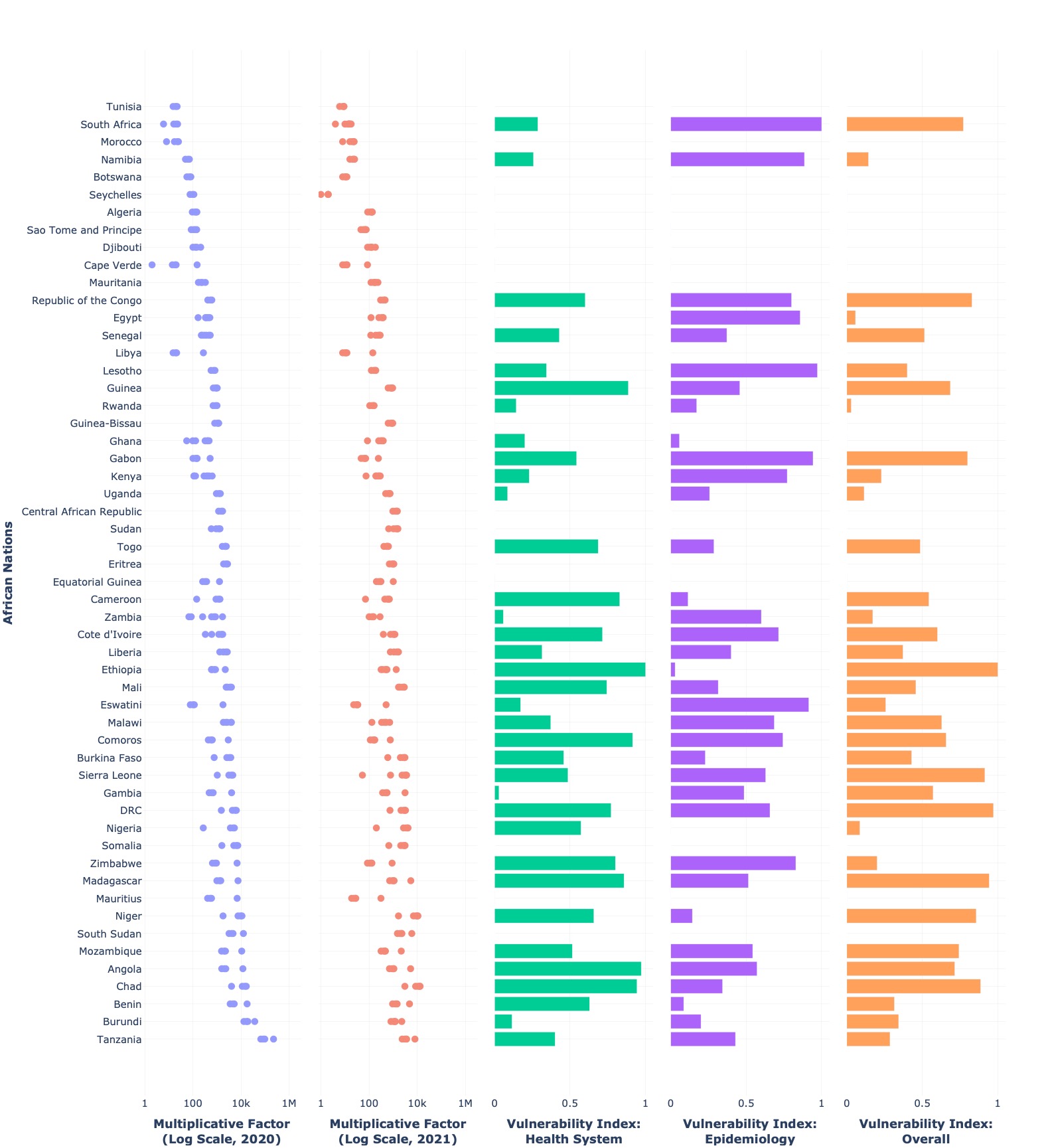

*A qualitative pan-method comparison of multiplicative factors and vulnerabilities. Rows with blanks for vulnerability denote unavailable data.*

**Supplementary Figure 4**

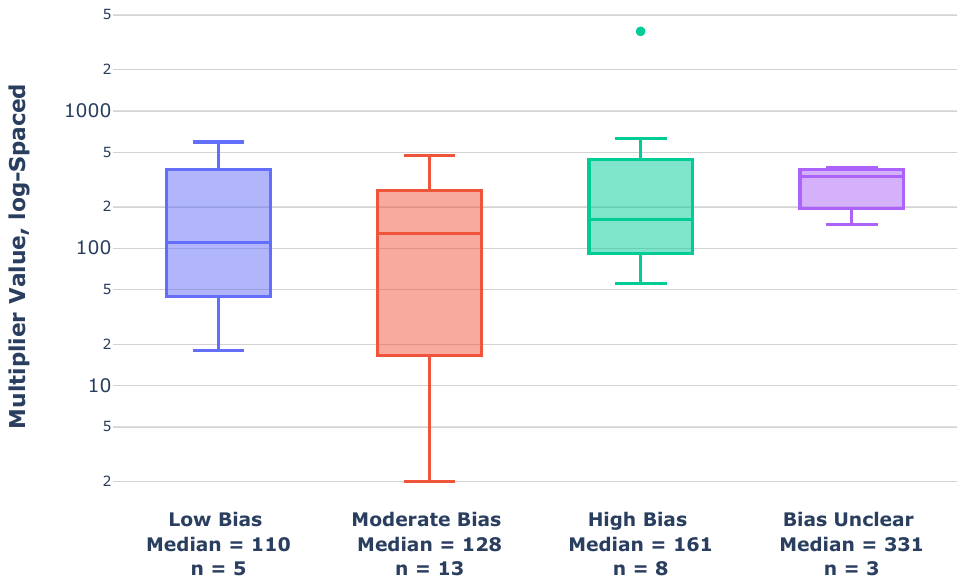

*Multiplicative factors compared to Joanna Briggs Institute-classified risk of bias (low, medium, high, or unknown).*

|  |  | **Zambia** | **South Africa** | **Ethiopia** | **Rwanda** |
| --- | --- | --- | --- | --- | --- |
| **2020** | **Projected Population (P20)** | 18,383,960 | 59,308,690 | 114,963,580 | 12,952,210 |
|  | **Crude All-Cause Mortality Rate Per 1,000 Population (CMR20)** | 6 | 9 | 6 | 5 |
|  | **Deaths (D20)**  **= P20 x CMR20** | 110,304 | 533,778 | 689,781 | 64,761 |
|  | **Zambia-Based Multiplier, Mwananyanda et al. (MULT20)** | 0.159 | 0.159 | 0.159 | 0.159 |
|  | **Expected Deaths With COVID-19 (ED20)**  **= D20 x MULT20** | 17,538 | 84,870 | 109,675 | 10,297 |
| **2021** | **Projected Population (P21)** | 18,920,657 | 60,041,996 | 117,876,226 | 13,276,517 |
|  | **Deaths (D21)**  **= P21 x CMR20** | 113,524 | 540,378 | 707,257 | 66,383 |
|  | **Zambia-Based Multiplier, Gill et al. (MULT21)** | 0.32 | 0.32 | 0.32 | 0.32 |
|  | **Expected Deaths With COVID-19 (ED21)**  **= D2021 x MULT21** | 36,328 | 172,921 | 226,322 | 21,242 |
|  | **April ’20 IFR, Sorensen et al.** | 0.14% | 0.51% | 0.15% | 0.18% |
|  | **Expected COVID-19 Infections Through 2021 (EC21)**  **= ED21 / April ’20 IFR** | 25,582,860 | 33,839,716 | 152,920,509 | 11,933,948 |
|  | **Adjusted Population**  **= Projected Population – D20** | 18,903,119 | 59,957,125 | 117,766,551 | 13,266,220 |
|  | **Adjusted Expected COVID-19 Infections Through 2021 (AEC21)**  **= ED21 / April ’20 IFR** | 25,559,146 | 33,791,883 | 152,778,228 | 11,924,692 |
|  | **% Difference**  **= (AEC21 – EC21) / EC21** | -0.09% | 0.14% | -0.09% | -0.08% |

**Supplementary Table 1: Sample Sensitivity Analysis for Infections, 2021 Population Size**
